# An Explainable AI-Driven Classification Framework for Parkinson’s Disease Detection via Acoustic Speech Features: A Comparative Machine Learning Study

**DOI:** 10.64898/2026.07.22.26358742

**Authors:** Dipraj Howlader, Tanvir Ahmed, Md. Mahbubur Rahman

## Abstract

Parkinson’s Disease (PD) is a progressive neurodegenerative disorder which significantly affects motor function, daily coordination and verbal communication. Speech-based biomarkers provide a non-invasive and scalable approach to early detection, as dysphonia is one of the earliest and most consistent clinical markers of PD. The dataset used in this study is publicly available and consists of 756 voice recordings from 252 subjects (188 with PD and 64 neurologically healthy controls) with a wide range of acoustic parameters such as Mel-Frequency Cepstral Coefficients (MFCCs), energy-based parameters, and higher-order statistical derivatives. After systematic preprocessing and z-score normalisation, five machine learning classifiers were tested: K-Nearest Neighbors (KNN), Extreme Gradient Boosting (XGBoost), Random Forest (RF), Support Vector Machine (SVM), and Naïve Bayes (NB) under a subject-independent, GroupKFold cross-validation protocol. The KNN classifier performed best overall with an accuracy of 92.10%, F1 score of 94.50%, and a precision rate of 98.09%, reducing the number of false positive diagnoses. To overcome the lack of interpretability of black-box predictive models, SHapley Additive exPlanations (SHAP) were used to explain the contribution of each feature to the prediction of an individual. The most diagnostically salient acoustic biomarkers were identified as features from the SHAP analysis: std_delta_delta_log_energy, the first Mel-Frequency Cepstral Coefficient, and Tunable Q-Factor Wavelet Transform (TQWT). This work introduces a machine learning framework that is both reproducible and clinically interpretable, combining high predictive accuracy with transparent, physiologically grounded decision logic.

## I. Introduction

PARKINSON’S Disease (PD) is the second most common neurodegenerative disorder in the world, after Alzheimer’s disease, and the burden of the disease has significantly increased in recent decades. Based on the global epidemiological data, in 2021, there were about 11.77 million people living with PD, which is a 274 percent increase from 3.15 million cases in 1990, with age-standardised prevalence at 138.63 per 100,000 persons [1], [5]. The cardinal motor symptoms of the disease, such as resting tremor, rigidity and bradykinesia, are usually observed only after significant dopaminergic neurodegeneration has taken place [2], [3]. The Braak hypothesis suggests that clinically apparent parkinsonism only occurs at stage three or four of Lewy body pathology, which involves the progressive accumulation of alpha-synuclein in the substantia nigra [4]. This neuropathological staging suggests that there is a pre-symptomatic period of 5-10 years before motor symptoms start, during which non-motor symptoms (NMS) can be identified [3].

In this pre-symptomatic stage, machine learning (ML) and deep learning (DL) techniques have shown promise in the detection of PD, even years before motor symptoms are clinically evident. One of the first and most consistent non-motor symptoms of PD is the acoustic irregularity of fundamental frequency, shimmer, jitter and spectral energy distribution, which are caused by the progressive loss of neuro-motor control of the larynx and respiratory system [6]. These measurable dysphonic features make speech signal analysis an ideal field for automated, non-invasive PD screening.

Although ML models have been shown to be accurate predictors, they are often criticized for their ‘black box’ quality, in which the logic behind a classification decision is opaque to both clinicians and patients [7], [8]. In clinical environments where stakes are high, the lack of interpretability makes it difficult to use AI tools responsibly for diagnosis, introduces medico-legal issues, and diminishes clinician confidence [9]. Explainable Artificial Intelligence (XAI) aims to fill this void by offering methods that help to understand the reasoning behind each model’s prediction. The SHapley Additive exPlanations (SHAP) framework, which is based on cooperative game theory, quantifies the contribution of each feature to a given prediction, allowing for both global feature importance ranking and localized, instance-level explanation [10].

The main goals of the present study are:

- To build and compare five ML classifiers for binary PD detection based on acoustic speech features;
- To use rigorous preprocessing, subject-level cross-validation and hold-out evaluation protocols to avoid data leakage and to ensure unbiased generalisation estimates;
- To interpret the best-performing model using SHAP analysis, identifying the most diagnostically informative vocal biomarkers; and
- To provide a reproducible, clinically transparent framework that connects algorithmic performance with medical inter-pretability.

## II. Literature Review

This section reviews three streams of previous research that have different thematic focus: speech-based ML methods for PD classification, multimodal diagnostic frameworks, and studies focused on XAI for clinical AI systems.

### A. Speech Based Machine Learning Approaches

There is an increasing amount of research that shows that voice recordings are a viable non-invasive diagnostic tool for PD. Soni et al. (2025) used a dataset of 756 voice recordings, with a train-test split of 80/20, and tested a Decision Tree, Gradient Boosting, and KNN classifier, using parameters such as Pitch Period Entropy (PPE), Detrended Fluctuation Analysis (DFA), and Recurrence Period Density Entropy (RPDE) [11]. The highest accuracy was obtained with the KNN with 88.8% which is better than Gradient Boosting (84.8%) and Decision Tree (78.2%) [11].

Bashir et al. (2023) showed that XGBoost with Recursive Feature Elimination (RFE) could reach 95% accuracy on a 195-instance dataset, but the authors noted that the small sample size limited the generalisability of their results [12]. A KNN model using perfect precision achieved an accuracy of 96.61% [13] and a Random Forest model with leave-one-out cross-validation achieved an accuracy of 99.94% [14]. A more comprehensive study of ML methods for prediction and progression of PD confirmed the overall performance of ensemble and distance-based classifiers over simpler probabilistic models [15].

### B. Multimodal Diagnostic Frameworks

The complementarity of heterogeneous data modalities for improving diagnostic precision has been explored recently. Rajeshpushpa et al. (2025) used a combination of wearable sensor time-series (from accelerometers and gyroscopes) and voice data from the UCI Parkinson’s Telemonitoring Dataset, which they processed using Convolutional Neural Networks (CNNs) with an overall accuracy of 97.3%, sensitivity of 96.8%, and specificity of 97.7% [16]. Tesfai (2023) used a multimodal ensemble model combining log-mel spectrograms and handcrafted acoustic features, achieving a classification accuracy of 99.82%, which is significantly higher than any single-modality pipeline [17]. A systematic review by Islam et al. (2024) also synthesized the evidence from both handwriting and voice datasets and showed that multimodal ensembles generally outperform unimodal classifiers in PD detection [18].

### C. Explainable AI in Clinical Applications

The use of XAI techniques in clinical ML systems has gained growing research interest. Alharthi et al. (2025) systematically reviewed 192 papers on nine popular XAI techniques, such as Layer-wise Relevance Propagation (LRP), LIME, SHAP, and Grad-CAM, for various sensor modalities including wearable inertial sensors, ECG, EEG, and neuroimaging [19]. They highlighted the importance of transparency in ML-driven sensor analysis for fostering the clinical trust and providing real-time decision support, yet also noted ongoing challenges such as the balance between model complexity and explainability, scalability limitations, and the requirement for the formats that are accessible to clinicians [19]. Shyamala and Navamani (2025) showed that the use of SHAP in a severity-stage classifier for PD significantly enhanced interpretability by quantifying the contribution of the individual speech features to the classification of a given class [20]. Together, these studies highlight the need to shift the focus from accuracy-based model evaluation to models that meet both performance and clinical transparency requirements.

One recurring shortcoming in the literature reviewed is the focus on model prediction over model explainability, and the evaluation on non-representative or imbalanced datasets. The present study directly tackles these gaps by combining stringent subject-independent validation with post-hoc interpretation using SHAP.

## III. Methodology

The methodology is sequential, consisting of data acquisition, data preprocessing, model training, subject-level evaluation, and explainability analysis. Particular focus is placed on methodological choices that guarantee statistical fairness, data leakage, and ethically appropriate diagnostic inference. The system is overall interpretable in terms of model behavior and subject-independent validation in the context of speech-based acoustic biomarkers, ensuring the statistical accuracy of the diagnostic predictions that are obtained, but also their clinical transparency and practical understandability. Figure 1 shows a schematic overview of the entire pipeline.

**Fig. 1.**
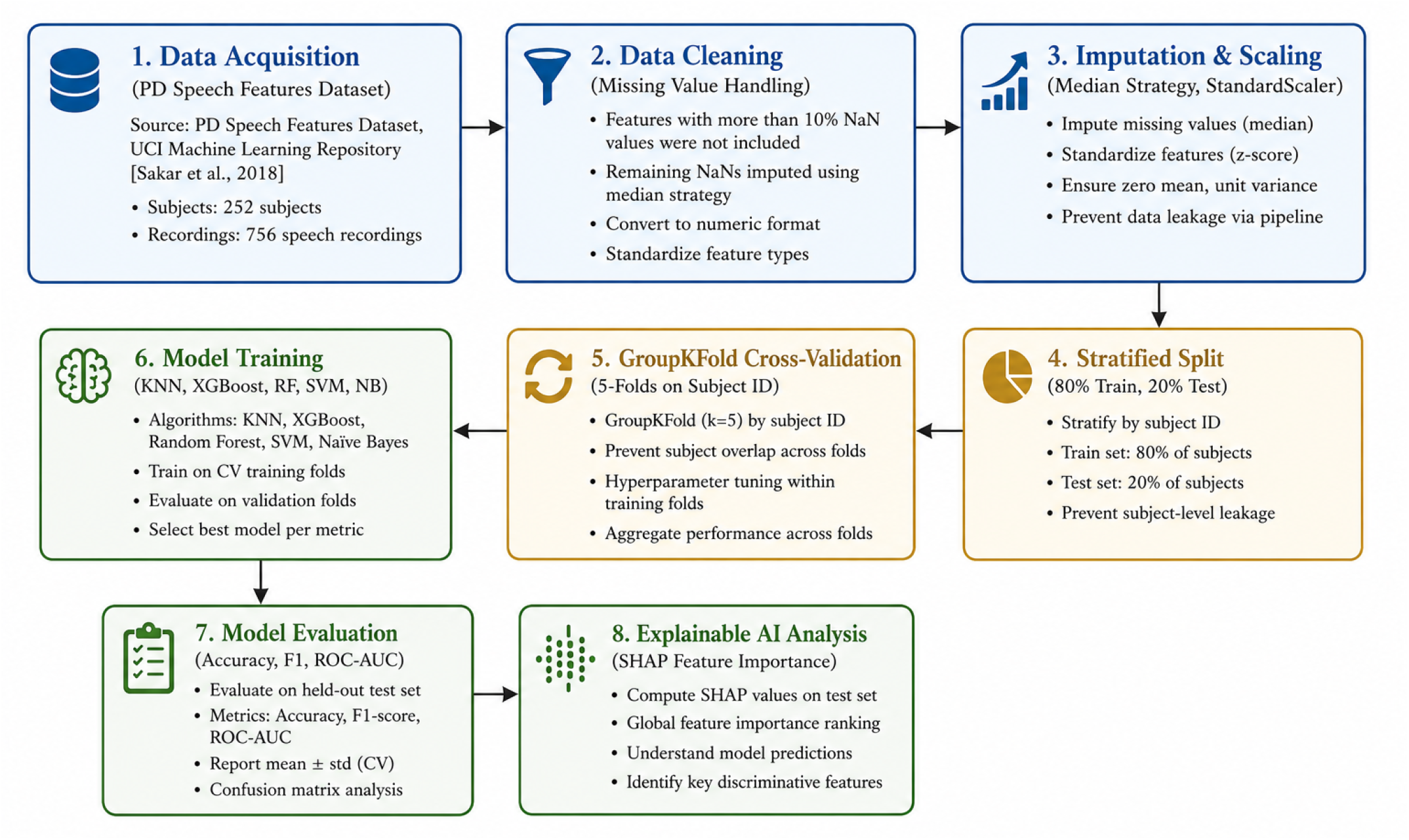
Overview of the machine learning pipeline for Parkinson’s disease detection using speech features. *Abbreviations: PD, Parkinson’s Disease; KNN, k-Nearest Neighbors; RF, Random Forest; SVM, Support Vector Machine; NB, Naïve Bayes; CV, Cross-Validation; SHAP, SHapley Additive exPlanations*.

### A. Dataset Description

The PD Speech Features Dataset [21], [22] is used in this study, which is publicly available and contains biomedical voice measurements recorded at the Department of Neurology, Cerrahpaşa Faculty of Medicine, Istanbul University. The raw data is a feature matrix of 755 features and 756 recordings (samples) from 252 subjects. The PD cohort consists of 188 patients (107 male, 81 female) with ages ranging from 33 to 87 years (mean ± SD: 65.1 ± 10.9), while the 64 healthy controls (23 male, 41 female) range in age from 41 to 82 years (61.1 ± 8.9). The entries contain quantitative acoustic features such as PPE, DFA, RPDE, MFCCs, and TQWT-derived features, as well as subject identification metadata.

### B. Data Preprocessing and Normalisation

Features with a missing value proportion greater than 10% were removed from the analysis to maintain the integrity of the feature matrix. Empirical inspection showed that there were no features removed at this threshold, which means that the data set was relatively complete. The missing values were imputed using the median strategy (SimpleImputer), which was preferred because it is less affected by outliers than the mean strategy. All features were then scaled using the StandardScaler (*µ*=0,*σ*=1), which was fitted only on the training data and applied to the test data to avoid data leakage.

### C. Train-Test Partitioning

Stratified sampling was used to split the preprocessed dataset into an 80% training set and a 20% held-out test set, with the same class proportions in both sets. To avoid subject-level data leakage, subject-level stratification was used, meaning that the same person’s recordings were not included in both splits.

### D. Classifier Definitions

The following five classifiers with different inductive biases were evaluated in comparison:

*1*) *K-Nearest Neighbors (KNN):* A non-parametric, lazy learning algorithm that classifies instances according to the proximity to the k = 5 training neighbours with the Euclidean distance metric [23].
*2*) *Random Forest (RF):* An ensemble of 100 decision trees using bagging to reduce variance and give intrinsic feature importance estimates [24].
*3*) *Support Vector Machine (SVM):* A maximum-margin classifier with a Radial Basis Function (RBF) kernel with probability outputs (probability = True) for threshold-based evaluation [25].
*4*) *Naïve Bayes (NB):* A probabilistic classifier based on the Bayes’ theorem with Gaussian likelihood distribution for each feature [26].
*5*) *XGBoost:* A regularised, sequential boosting ensemble with hyperparameters n estimators = 100, max depth = 3, learning rate = 0.2, and others [27].

### E. Performance Metrics

The performance of the models was measured using five standard classification metrics:

*1*) *Accuracy:* The overall proportion of correct predictions;
*2*) *Precision:* The ratio of true positives to all predicted positives (TP / (TP + FP));
*3*) *Recall (Sensitivity):* The ratio of true positives to all actual positives (TP / (TP + FN)), reflecting the model’s capacity to avoid missed diagnoses;
*4*) *F1-Score:* The harmonic mean of precision and recall, providing a balanced composite measure; and
*5*) *ROC-AUC:* The area under the Receiver Operating Characteristic curve, quantifying discriminative ability across classification thresholds.

### F. Subject-Level Cross-Validation

To avoid the leakage of subject-level information in the training process, a 5-fold GroupKFold cross-validation approach was used, where folds were grouped by the subject identifier [28]. This technique guarantees that every recording for every subject is part of a single fold (either train or test). GroupKFold was applied within the 80% training set; the 20% test set remained held out throughout.

*1*) *True Label:* The aggregated subject label (y true) was set to the maximum class value across all of a subject’s recordings. This means that if any recording for a subject was labeled as PD (1), the subject’s overall true label was 1.
*2*) *Prediction Score:* The subject prediction score (y prob) was calculated as the mean probability of the positive class across all of the subject’s recordings.

### G. Explainability via SHAP

The post-hoc model interpretation was performed with the SHapley Additive exPlanations (SHAP) framework, which uses cooperative game theory to calculate a contribution value for each feature [10]. The best-performing KNN model on the held-out test set was then used with the PermutationExplainer to obtain global feature importance rankings and localised, instance-level explanation vectors. SHAP values allow clinical practitioners to gain insight into which acoustic properties are most significantly contributing to individual predictions, thus providing a physiologically meaningful biomarker evidence to support algorithmic outputs.

## IV. Results

A detailed performance comparison is given in Table I for all the five classifiers. The KNN model had the best overall performance with an accuracy of 92.10%, an F1 score of 94.50%, a precision of 98.09% and an ROC-AUC of 94.21%. Higher precision suggests that the false-positive rate is very low, especially in the clinical setting where the test is used for diagnostic screening to prevent unnecessary burden on patients.

**TABLE I.** Performance Comparison of Different Machine Learning Models.

| Model | Accuracy (%) | Precision (%) | Recall (%) | F1-Score (%) | ROC-AUC (%) |
| --- | --- | --- | --- | --- | --- |
| KNN | 92.10 | 98.09 | 91.15 | 94.50 | 94.21 |
| XGBoost | 86.84 | 87.80 | 95.57 | 91.53 | 92.74 |
| Random Forest | 84.87 | 84.62 | 97.35 | 90.54 | 91.62 |
| SVM | 82.24 | 82.58 | 96.46 | 88.98 | 87.66 |
| Naïve Bayes | 73.68 | 85.44 | 77.88 | 81.48 | 72.34 |

XGBoost had the second highest recall (95.57%) after Random Forest (97.35%), which had the highest recall of all classifiers, a clinically significant advantage as there is a strong need to reduce the number of false negatives in PD screening. The accuracy of XGBoost was 86.84% and the F1 score was 91.53%. The Random Forest classifier was the second best with 84.87% accuracy and 91.62% ROC-AUC, and it had a good balance of sensitivity and overall discriminative ability. The SVM classifier performed with an accuracy of 82.24% and an F1-score of 88.98%, which is reasonable but not as good as the ensemble-based classifiers in terms of accuracy and discriminative power. The Naïve Bayes classifier achieved the lowest accuracy (73.68%) and ROC-AUC (72.34%) as it assumes that the features are conditionally independent, which is not well suited to the highly correlated acoustic feature space of speech-based PD datasets. Figure 2 illustrates the model comparison plot.

**Fig. 2.**
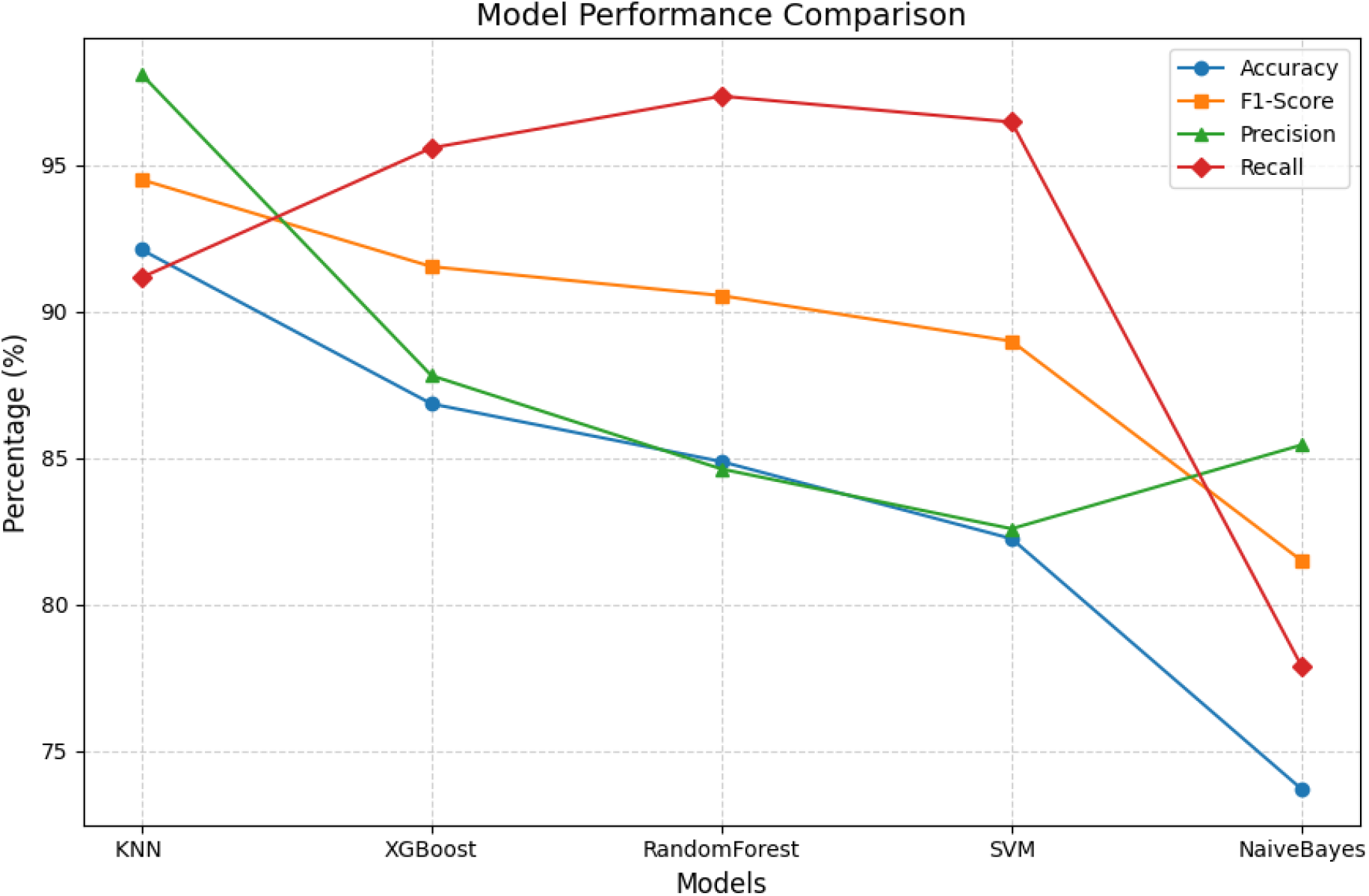
Performance comparison of KNN, XGBoost, Random Forest, SVM, and Naïve Bayes using Accuracy, F1-score, Precision, and Recall.

### A. SHAP Explainability Analysis

To understand the contribution of individual features to each prediction, the best performing KNN model was subjected to SHAP analysis. Figure 3 shows the SHAP summary plot, where each point represents the SHAP value of a feature for a single observation; the further to the right a point is, the higher the probability of a PD classification, and the further to the left a point is, the higher the probability of a healthy classification. Four feature clusters were found to be the most diagnostic:

**Fig. 3.**
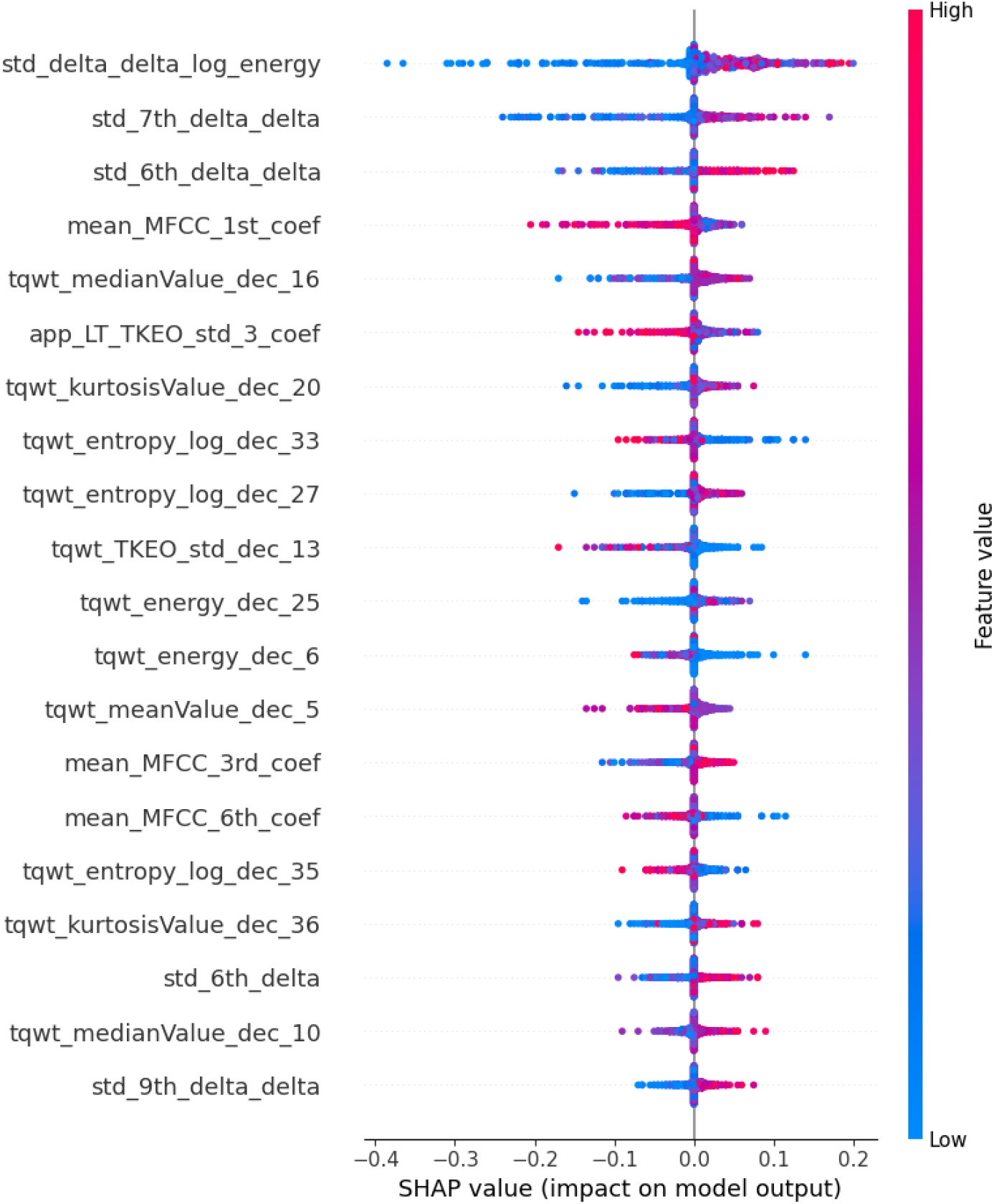
SHAP summary plot showing the importance and contribution of the top speech features to Parkinson’s disease classification.

*1*) *std delta delta log energy:* The feature with the highest mean SHAP magnitude was std delta delta log energy, which represents second-order temporal changes in log-energy of the voice. This salience supports clinical observations that PD patients have reduced control over the strength of their voice and the support of their breath, which leads to irregular energy dynamics between speech segments.
*2*) *mean MFCC 1st coef:* The first Mel-Frequency Cepstral Coefficient is the spectral envelope of the speech at low frequencies. This high SHAP value is in line with previous reports of spectral flattening in PD patients, which is due to decreased articulatory effort and laryngeal motor control.
*3*) *TQWT-derived features (entropy, energy, kurtosis, median):* The features derived from the Tunable Q-Factor Wavelet Transform (TQWT) at different decomposition levels were found to be significant contributors together. The TQWT’s ability to resolve oscillatory components at the programmable frequency resolutions renders it especially sensitive to the small aperiodic variations in the voice that are typical of PD-related dysphonia.
*4*) *app LT TKEO std 3 coef:* Coefficients from the Teager–Kaiser Energy Operator (TKEO) contributed moderately and consistently to model predictions. The TKEO is sensitive to instantaneous energy changes in quasi-periodic signals, indicating that rapid energy modulations in the phonation signal contain meaningful diagnostic information.

## V. Discussion

The results of this study confirm the feasibility of speech-based machine learning for non-invasive PD classification, and the accuracy (92.10%) and precision (98.09%) achieved by KNN are comparable and, in some aspects, superior to the accuracy and precision reported in similar previous studies based on the same datasets [11], [13]. The extremely high accuracy of the KNN model has a specific clinical relevance: a false positive in a PD screening context means that the patient will undergo further diagnostic evaluation, causing anxiety and potentially over-treatment, with a high cost to the patient and health care systems. In contrast, the best recall was seen with Random Forest (97.35%) which would be desirable in screening situations where the goal is to reduce missed diagnoses while increasing false positive rates.

Ensemble methods (Random Forest, XGBoost) were the most dominant in terms of recall, while KNN outperformed the others in terms of accuracy and precision, indicating that there is no single best classifier for all clinical goals. Instead, the choice of classifiers should depend on the particular decision problem, such as whether it is more important to minimize the number of missed cases (recall) or the number of unnecessary referrals (precision).

SHAP analysis offers a clinically relevant level of interpretability in addition to classification metrics. The most influential feature identified (std delta delta log energy) is consistent with known neurophysiology of dysphonia in PD: the basal ganglia are known to be involved in dysphonia in PD, and the voluntary modulation of respiratory and laryngeal motor control is impaired, leading to irregular patterns of vocal energy. Likewise, the importance of MFCC features and TQWT-derived coefficients is in line with previous spectral and wavelet studies of PD speech, which further supports the diagnostic value of these acoustic features [6], [21].

## VI. Conclusion

In this study, a rigorous and interpretable machine learning system for detecting Parkinson’s Disease based on acoustic speech features was proposed. In the overall balance of accuracy (92.10%), precision (98.09%), and F1-score (94.50%), KNN was the most reliable model for clinical PD screening in this dataset among five evaluated classifiers. SHAP-based post-hoc explainability analysis revealed that second-order temporal energy dynamics (std delta delta log energy), low-frequency spectral envelope coefficients (mean MFCC 1st coef), and multi-resolution wavelet energy features (TQWT) constitute the most diagnostically informative acoustic biomarkers. The results are physiologically sound and build on previous research on vocal dysfunction in PD.

There are some limitations to the generalisability of the present results. The data was gathered in a controlled clinical setting with one institution, which restricts ecological validity in heterogeneous recording contexts, languages, and demographic populations. To overcome these limitations, future research is needed to build naturally interpretable or hybrid architectures that integrate explainability into the training process, integrate multimodal physiological signals (e.g., handwriting kinematics, gait dynamics, facial expression analysis, or brain neuroimaging), and perform prospective clinical validation in collaboration with neurologists and speech-language pathologists. The conversion of this framework to a user-friendly diagnostic tool, like a mobile or web application, would also broaden its clinical usefulness and make it easier to conduct large-scale, community-based early screening.

However, the present work offers a methodologically sound and meaningful basis for automated, fair PD detection even with limited voice data. The ability to predict accurately and be transparent about the predictions using SHAP analysis is a great example of how responsible AI can be used in clinical diagnostics and help realize the vision of intelligent, inclusive, and accountable healthcare systems.

## Data Availability

The data used in this study are publicly available and were obtained from the original public repository. The dataset can be accessed at: https://archive.ics.uci.edu/dataset/470/parkinson+s+disease+classification. No new data were generated during this study.

## References

[1] Y. Luo, L. Qiao, M. Li, X. Wen, W. Zhang, and X. Li, “Global, regional, national epidemiology and trends of Parkinson’s disease from 1990 to 2021: findings from the Global Burden of Disease Study 2021,” Frontiers in Aging Neuroscience, vol. 16, p. 1498756, 2025.

[2] GBD 2021 Parkinson’s Disease Collaborators, “Projections for prevalence of Parkinson’s disease and its driving factors in 195 countries and territories to 2050: modelling study of Global Burden of Disease Study 2021,” BMJ, vol. 388, p. e080952, 2025.

[3] A. H. V. Schapira, K. R. Chaudhuri, and P. Jenner, “Non-motor features of Parkinson disease,” Nature Reviews Neuroscience, vol. 18, no. 7, pp. 435–450, 2017.

[4] H. Braak, K. Del Tredici, U. Rüb, R. A. I. de Vos, E. N. H. Jansen Steur, and E. Braak, “Staging of brain pathology related to sporadic Parkinson’s disease,” Neurobiology of Aging, vol. 24, no. 2, pp. 197–211, 2003.

[5] M. Li, X. Ye, Z. Huang, L. Ye, and C. Chen, “Global burden of Parkinson’s disease from 1990 to 2021: a population-based study,” BMJ Open, vol. 15, no. 4, p. e095610, 2025.

[6] A. Tsanas, M. A. Little, P. E. McSharry, and L. O. Ramig, “Accurate telemonitoring of Parkinson’s disease progression by noninvasive speech tests,” IEEE Transactions on Biomedical Engineering, vol. 57, no. 4, pp. 884–893, 2010.

[7] P. Linardatos, V. Papastefanopoulos, and S. Kotsiantis, “Explainable AI: a review of machine learning interpretability methods,” Entropy, vol. 23, no. 1, p. 18, 2020.

[8] A. Adadi and M. Berrada, “Peeking inside the black-box: a survey on explainable artificial intelligence (XAI),” IEEE Access, vol. 6, pp. 52138–52160, 2018.

[9] C. Rudin, “Stop explaining black box machine learning models for high stakes decisions and use interpretable models instead,” Nature Machine Intelligence, vol. 1, no. 5, pp. 206–215, 2019.

[10] S. M. Lundberg and S.-I. Lee, “A unified approach to interpreting model predictions,” in Proc. 31st Conference on Neural Information Processing Systems (NeurIPS), vol. 30, 2017, pp. 4765–4774.

[11] T. Soni, D. Gupta, M. Dutta, B. V. Kumar, and H. MuhamedAle, “Speech-based Parkinson’s disease classification with KNN: achieving high accuracy in non-invasive diagnostics,” in Proc. 2025 4th OPJU International Technology Conference (OTCON), Raigarh, India, 2025, pp. 1–4.

[12] A. Bashir, Y. Singh, S. Zadoo, and K. H. Mir, “Parkinson’s disease detection: a machine learning-based model,” in Proc. 6th International Conference on Futuristic Trends in Networks and Computing Technologies (FTNCT), Chandigarh, India, Jul. 2023, Lecture Notes in Electrical Engineering, Springer.

[13] Rhythm R. Khandelwal, P. Badlani, and B. Saxena, “Dysphonic voice pattern-based Parkinson disease detection using machine learning models,” in Proc. 2024 2nd International Conference on Disruptive Technologies (ICDT), Greater Noida, India, 2024, pp. 1475–1479.

[14] V. Shibina and T. M. Thasleema, “Acoustic signal-based diagnosis of neurodegenerative Parkinson’s disease through a machine learning approach: a review,” in Proc. 2022 IEEE 19th India Council International Conference (INDICON), Kochi, India, 2022, pp. 1–8.

[15] S. Gaba and H. Kaur, “Machine learning techniques for Parkinson’s disease prediction and progression: a comprehensive review,” in Proc. 2024 International Conference on Communication, Computer Sciences and Engineering (IC3SE), Gautam Buddha Nagar, India, 2024, pp. 430–436.

[16] J. Rajeshpushpa, N. Radha, and L. J. Mary, “Speech-based early detection of Parkinson’s disease,” in Proc. 2025 International Conference on Innovative Trends in Information Technology (ICITIIT), Kottayam, India, 2025, pp. 1–6.

[17] S. Tesfai, “Multimodal ensemble models for Parkinson’s disease diagnosis using log-mel spectrograms and acoustic features,” in Proc. 2023 IEEE MIT Undergraduate Research Technology Conference (URTC), Cambridge, MA, USA, 2023, pp. 1–5.

[18] M. A. Islam, M. Z. H. Majumder, M. A. Hussein, K. M. Hossain, and M. S. Miah, “A review of machine learning and deep learning algorithms for Parkinson’s disease detection using handwriting and voice datasets,” Heliyon, vol. 10, no. 3, p. e25469, 2024.

[19] A. S. Alharthi et al., “Explainable AI for sensor signal interpretation to revolutionize human health monitoring: a review,” IEEE Access, vol. 13, pp. 115990–116024, 2025.

[20] K. Shyamala and T. M. Navamani, “Design of an optimized feature-driven severity stage classifier for Parkinson’s disease prediction using deep learning,” IEEE Access, vol. 13, pp. 142140–142160, 2025.

[21] C. O. Sakar, G. Serbes, A. Gunduz, H. C. Tunc, H. Nizam, B. E. Sakar, M. Tutuncu, T. Aydin, M. E. Isenkul, and H. Apaydin, “A comparative analysis of speech signal processing algorithms for Parkinson’s disease classification and the use of the Tunable Q-Factor Wavelet Transform,” Applied Soft Computing, vol. 74, pp. 255–263, 2019.

[22] M. A. Little, P. E. McSharry, and S. J. Roberts, “Exploiting nonlinear recurrence and fractal scaling properties for voice disorder detection,” BioMedical Engineering OnLine, vol. 6, no. 1, p. 23, 2007.

[23] T. Cover and P. Hart, “Nearest neighbor pattern classification,” IEEE Transactions on Information Theory, vol. 13, no. 1, pp. 21–27, 1967.

[24] L. Breiman, “Random forests,” Machine Learning, vol. 45, pp. 5–32, 2001.

[25] C. Cortes and V. Vapnik, “Support-vector networks,” Machine Learning, vol. 20, pp. 273–297, 1995.

[26] R. O. Duda, P. E. Hart, and D. G. Stork, Pattern Classification, 2nd ed. New York, NY, USA: Wiley-Interscience, 2001.

[27] T. Chen and C. Guestrin, “XGBoost: a scalable tree boosting system,” in Proc. ACM SIGKDD International Conference on Knowledge Discovery and Data Mining (KDD), San Francisco, CA, USA, 2016, pp. 785–794.

[28] F. Pedregosa et al., “Scikit-learn: machine learning in Python,” Journal of Machine Learning Research, vol. 12, pp. 2825–2830, 2011.

